# Avoiding versus contracting COVID-19: On the effectiveness of social distancing at the level of the individual

**DOI:** 10.1101/2020.10.29.20222422

**Authors:** Russell H. Fazio, Benjamin C. Ruisch, Courtney A. Moore, Javier A. Granados Samayoa, Shelby T. Boggs, Jesse T. Ladanyi

## Abstract

Past research has established the value of social distancing as a means of deterring the spread of COVID-19 largely by examining aggregate level data. Locales in which efforts were undertaken to encourage distancing experienced reductions in their rate of transmission. However, these aggregate results tell us little about the effectiveness of social distancing at the level of the individual, which is the question addressed by the current research. Four months after participating in a study assessing their social distancing behavior, 2,120 participants indicated whether they had contracted COVID-19. Importantly, the assessment of social distancing involved not only a self-report measure of how strictly participants had followed social distancing recommendations, but also a series of virtual behavior measures of social distancing. These simulations presented participants with graphical depictions mirroring specific real-world scenarios, asking them to position themselves in relation to others in the scene. Individuals’ social distancing behavior, particularly as assessed by the virtual behavior measure, predicted whether they contracted COVID-19 during the intervening four months. This was true when considering only participants who reported having tested positively for the virus and when considering additional participants who, although untested, believed that they had contracted the virus. The findings offer a unique form of additional evidence as to why individuals should practice social distancing. What the individual does matters, not only for the health of the collective, but also for the specific individual.

---

Since its emergence in late 2019, the COVID-19 pandemic has had a devastating impact on nations around the world. The negative effects are evident with respect to the shocking number of people who have been infected with this novel coronavirus and the sheer number of deaths, as well as the impact upon nations’ economies. Given the current lack of a vaccine, minimizing the spread of COVID-19 has required that people change their behavior. Government officials and public health experts worldwide have urged people to engage in various preventive actions: wash your hands frequently for at least 20 seconds, use hand sanitizer when handwashing is not feasible, avoid touching your face, disinfect surfaces that you touch, and wear masks. Perhaps most importantly, they have pleaded that their citizens engage in social distancing – avoid close contact with other people. The mantra “six feet” has been repeated regularly and now appears on signs and floor decals throughout society.

Although knowledge regarding SARS-CoV-2 transmission is constantly expanding, much has been learned about the dynamics responsible for its spread (Lee, Wada, Grabowski, Gurley, & Lessler, 2020). Considerable evidence stemming from the investigation of such viruses as SARS, MERS, and SARS-CoV-2 highlights that physical distancing reduces their aerial transmission (e.g., Chu et al., 2020). Mathematical models of the spread of COVID-19 in particular (e.g., Ferguson et al., 2020; Greenstone & Nigam, 2020; Kissler, Tedijanto, Lipsitch, & Grad, 2020) have illustrated the efficacy of social distancing as a means of mitigating its spread. Moreover, recent epidemiological evidence also documents the effectiveness of social distancing at the societal level. For example, a recent analysis involving both the US states and 134 nations revealed that the implementation of social distancing policies was associated with a reduction in COVID-19 spread rates within those locales (McGrail, Dai, McAndrews, & Kalluri, 2020). Similarly, an examination of the daily growth rate in COVID-19 cases across US counties during March and April of 2020 found that the imposition of shelter-in-place orders and closures of restaurants, bars, and entertainment-related businesses slowed the spread of COVID-19 (Courtemanche, Garuccio, Le, Pinkston, & Yelowitz, 2020). Other analyses have found that greater social distancing at the county level, as estimated by smartphone GPS data, is associated with reduced spread of COVID-19 and fewer deaths (VoPham et al., 2020).

Yet what remains unknown is the extent to which an *individual’s* social distancing behavior predicts their personal likelihood of contracting COVID-19. This is a critical theoretical and empirical gap, particularly given that the demonstrated effectiveness of social distancing at the aggregate level does not necessarily mean that individual differences in social distancing behavior will predict whether a given individual contracts the disease. For example, it may be that a “critical mass” of social distancing behavior in a given locale is necessary for any individual-level benefits to emerge. Until that critical level is reached, it is possible that individual differences may matter little for disease transmission.

Furthermore, beyond the question of *whether* individual differences in social distancing behavior predict disease transmission, there is also the critical question of the *degree* to which these individual differences matter. In other words, what is the size of this effect? Do those who practice social distancing enjoy a notably reduced likelihood of contracting the disease? Or are these effects relatively modest in size? Aggregate data offer little insight into these questions. Indeed, aggregation over a large number of instances may be necessary to observe what might be small effects at the level of the individual.

To address this theoretical and empirical gap, the current research pursues a different approach to testing the effectiveness of social distancing – one that focuses not on aggregates but on the level of the individual. Are individuals who engage in social distancing behaviors less likely to contract COVID-19? If so, to what degree? To examine this question, we assessed participants’ (US residents) social distancing behavior at the height of the first wave of the pandemic, and then four months later asked them to report whether they had contracted the virus.

The assessment of the extent to which individuals practiced social distancing was accomplished by two very different measurement techniques. The standard approach generally employed in social science research, and certainly true of hundreds of recent studies concerning the pandemic (Gollwitzer, Martel, Marshall, Höhs, & Bargh, 2020), is simply to prepare survey questions asking people to report the extent to which they personally practice social distancing. We did precisely the same in the current research.

However, what makes our research all the more unique is that we supplemented the self-report measure with a more innovative, behaviorally-oriented approach to the assessment of social distancing. We simulated social distancing behavior by presenting participants with graphical depictions that mirrored different real-world scenarios and asking them to position themselves in relation to others in the scene. These virtual social distancing scenarios required a concrete, “in-the-moment” behavioral decision, which could vary in the degree to which participants did or did not distance themselves from others. For example, in one scenario participants chose whether to cross a park via a circuitous but isolated path versus a more-direct but crowded route. In another, they were presented an aerial image of a crowded beach and asked to click on the spot where they personally would lay down their towel. Yet another presented an interactive image of two people approaching each other in a crosswalk for which participants were asked to move a slider that shifted the walkers from the center of the crosswalk to the distance that they personally would leave between themselves and the other individual. The ten behavioral scenarios can be viewed at our demonstration website, http://psychvault.org/social-distancing-measures/.

We included this virtual behavior distancing measure out of concern for the potential problems that have been documented to be associated with self-reports of behavior. Individuals may over-report their social distancing to convey a socially desirable impression to others and themselves (Balcetis, 2008; Fisher, 1993; Gur & Sackeim, 1979; Leary & Kowalski, 1990). Moreover, self-reports may be all the more problematic to the extent that they rely on retrospective memory regarding past behavior (Kouchaki & Gino, 2016; Ross, 1989). By simulating concrete real-world situations that require an immediate decision, the graphical scenarios offer a means, in addition to a self-report, of indexing the extent to which individuals behave in a manner that accords with the principle of social distancing.

The present study examines whether both the self-report measure and the virtual behavior measure of social distancing prospectively predict, four months later, whether an individual contracted COVID-19. The data are available on the Open Science Framework: https://osf.io/x79ak/?view_only=b380d69aedc34bb98cf2670c04ecaa20.

## Method

We recruited our participant samples from Amazon’s Mechanical Turk participants (see Buhrmester, Kwang, & Gosling, 2011). Although not representative of the U.S. population, MTurk samples tend to be more demographically, politically, and geographically diverse than the samples typically used in psychological research (Paolacci & Chandler, 2014). They also perform similarly to non-MTurk samples across many tasks and measures (Berinsky, Huber, & Lenz, 2012; Hauser, Paolacci, & Chandler, 2019), including surveys on political attitudes (Clifford, Jewell, & Waggoner, 2015). Most importantly, however, our aim is not to make claims regarding the absolute frequency of social distancing behaviors or COVID-19 illness. Instead, we seek to understand whether the extent to which individuals engage in social distancing, as measured at one point in time, relates to their having contracted the coronavirus, as reported at a later point in time. Given this covariation aim, MTurk participants offer the opportunity for an appropriate test of the hypothesis that social distancing behavior is prospectively predictive of individuals’ reports of having become ill with COVID-19.

### Participants

The sample consisted of MTurkers who had participated in one of two earlier studies conducted in Spring 2020. All participants who had granted permission to re-contact them were invited to complete a brief survey approximately four months after their initial study for a payment of $1. A total of 2,120 individuals, all US residents, did so (1,031 women, 1,074 men, 15 no response; *M*_*age*_ = 40.39, *SD*_*age*_ = 15.34). Study 1 had been completed on May 7-8 (*n* = 1281) and Study 2 on June 9 (*n* = 839).^1^

### Measures

Each of the initial studies concerned reactions to the pandemic and relevant individual difference measures. The variables of interest for the current research concern social distancing behavior and two variables identified on an a priori basis as likely predictors of contracting COVID-19: preexisting conditions and working outside the home. The former was assessed by asking participants to consider their “personal health prior to the outbreak of the COVID-19 virus” and to then indicate whether they would have described themselves “as having pre-existing medical conditions that left you more vulnerable to the virus than the average person” by selecting one of five response options: Definitely not/ Probably not/ Maybe/ Probably yes/ Definitely yes (coded as 1-5, respectively). Participants were also asked about their employment status. Our interest was in whether they selected the option “My job requires that I leave my home” (coded as 1) as opposed to other options (coded as 0) concerning working from home, having lost their job due to COVID-19, or having been unemployed before the pandemic began. Our reasoning was that individuals whose job required leaving home would be at greater risk for contracting the virus.

#### Self-reported social distancing behavior

Participants also were asked to report on the extent to which they were practicing social distancing. For Study 1, which was conducted when only a few states had even begun the process of relaxing stay-at-home orders and re-opening their economies, a single relevant question was employed. Participants were asked “Generally speaking, how strictly have you personally been following the ‘social distancing’ recommendations?” and responded using a 7-point scale ranging from “not at all” to “very strictly.” By the time of Study 2, most states had re-opened, at least partially. Hence, we included two relevant questions involving the same 7-point scale. One inquired specifically about behavior during the lockdown: “Up to a few weeks ago, most people were under ‘shelter-in-place’ orders (for example, staying home except for absolute necessities, having no contact with people outside your household, etc.). During that time, how strictly did you follow these recommendations?” A second question asked specifically about maintaining safe distances: “Generally speaking, how strictly have you personally been following the “social distancing” recommendations of the government and CDC to maintain a distance of six feet or more from others?” Responses to these two questions correlated highly (*r* = .73) and, hence, were averaged to form a single composite of self-reported social distancing for the participants in this later study.

#### Virtual social distancing behavior

Following the provision of informed consent, each of the initial studies began with the presentation of graphical scenarios that asked participants to position themselves relative to others in a specific real-world situation. A total of ten behavioral scenarios, all similar to the examples described earlier, were presented. The scenarios are described in the Supplemental Material and, as noted earlier, can be viewed at our demonstration website, http://psychvault.org/social-distancing-measures/. After standardizing scores from each scenario within each study, we computed the average as our index of virtual social distancing behavior (α = .83).

#### Follow-up survey

Four months after the initial study, the participants completed a brief survey about whether they had or had not contracted the coronavirus. They first were asked whether they had been tested for COVID-19. If so, they indicated whether the test showed that they had COVID-19. If they had not been tested, they were asked “Even though you may not have been tested, do you believe that you have ever had COVID-19 / the coronavirus?” to which they responded yes or no. (These very same questions were included in the initial survey that participants completed.) Participants who reported either testing positively or believing they had COVID-19 were then asked to select from a list of possibilities how they thought they might have contracted the virus.

## Results

### Descriptive Data

Of the 2,120 participants, 516 (24.3%) reported having been tested for COVID-19, with 116 (5.5% of the total sample) reporting a positive result. Another 232 participants (10.9% of the total sample) reported that, although they had not been tested, they believed that they had indeed contracted the coronavirus. In sum, at the time of the follow-up survey, a total of 348 participants (16.42%) reported having experienced COVID-19 illness. When asked to indicate how they might have contracted the virus, 23.3% of these 348 participants selected “from a member of my household,” 21.9% selected “at work,” and 17.6% selected “at an indoor setting (a restaurant, bar, store, church or other indoor venue).” The other possibilities that were selected with some frequency were: “from a friend or relative who visited me at home” (11.2%), “at an outdoor setting (a public event, park, restaurant patio, outdoor party, etc.)” (6.9%), and “while traveling on a trip out of town” (4.9%).

### Predicting COVID-19 Illness at Follow-up

Our major interest is to examine whether social distancing behavior *prospectively predicts* subsequent illness. Hence, we need to exclude from the analyses any participants who reported having had COVID-19 at the time of their initial study. Of the 2,120 follow-up participants, 235 reported a positive test result at Time 1 or the untested belief that they had COVID-19, resulting in a sample of 1885 participants for our first set of analyses. Of these, 199 subsequently reported a positive test result or the belief that they had contracted COVID-19 (coded as 1) and 1,686 who reported either a negative test result or the belief that they had not contracted the virus^2^ (coded as 0).

A binary logistic regression was conducted examining the dichotomous COVID-19 status variable at follow-up as a function of the predictors variables of interest. To ease interpretation, all continuous predictor variables were standardized. We first considered the two a priori risk variables of preexisting conditions and leaving home for work (coded 0/1 for no/yes). As anticipated, each accounted for significant unique variance (pre-existing conditions: *B* = .36, Wald = 27.03, *p* < .001, odds ratio = 1.44; leave home for work: *B* = .41, Wald = 5.48, *p* = .019, odds ratio = 1.51). We next considered the virtual behavioral and self-report measures of social distancing, which correlated at .50, *p* < .001. When the behavioral distancing score was added as the next variable in the equation, it too revealed a statistically significant effect (*B* = -.22, Wald = 8.25, *p* = .004 odds ratio = .80); less social distancing was associated with a greater likelihood of contracting COVID-19. A similar effect, albeit weaker, was observed when the self-report measure of social distancing was added as the third variable in the equation, instead of the virtual behavior measure (*B* = -.13, Wald = 3.04, *p* = .081, odds ratio = .88). Table 1 presents the relevant statistics when all four variables are considered simultaneously. The virtual behavior measure accounted for significant unique variance, whereas the self-report measure did not.

**Table 1.** Binary Logistic Regression Predicting COVID-19 Status^a^.

| Variable | *B* | Standard Error | Wald | *p* | Odds ratio |
| --- | --- | --- | --- | --- | --- |
| Preexisting Conditions<sup>b</sup> | .395 | .072 | 30.326 | .000 | 1.484 |
| Leave Home for Work<sup>a</sup> | .350 | .183 | 3.684 | .055 | 1.420 |
| Self-Reported Behavior<sup>b</sup> | -.031 | .085 | .131 | .717 | .970 |
| Virtual Behavior<sup>b</sup> | -.202 | .087 | 5.459 | .019 | .817 |
| Constant | -2.286 | .091 | 624.055 | .000 | NA |

<sup>a</sup>Coded as 0=No/1=Yes; <sup>b</sup>Standardized

Parallel analyses were conducted predicting whether participants reported having tested positively for COVID-19 at follow-up. In other words, the focus here is solely on positive test results as an indicator of COVID-19 illness, in contrast to the earlier approach in which participants who had not been tested but believed they had contracted the virus also were considered within the COVID-19 classification. These analyses excluded only participants who had reported a positive test result in their initial survey. They compare the 85 participants who subsequently tested positively to the 1,993 who reported either a negative test or not having been tested at all.^3^ As before, all continuous predictor variables were standardized and we first considered the two a priori risk variables of preexisting conditions and leaving home for work. The former accounted for substantial unique variance (*B* = .69, Wald = 44.14, *p*<.001, odds ratio = 1.98). Whether participants worked away from home achieved a more marginal level of statistical significance (*B* = .43, Wald = 2.64, *p* = .104, odds ratio = 1.53). When entered next, the virtual behavior measure was, as before, statistically significant (*B* = -.22, Wald = 3.95, *p* = .047, odds ratio = .80), with less social distancing being associated with a greater likelihood of testing positively for COVID-19. This was not true when the self-report measure of social distancing was instead entered as the third variable in the equation (*B* = -.09, Wald = .53, *p* = .466, odds ratio = .92). Table 2 presents the relevant statistics when all four variables are considered simultaneously. Once again, the virtual behavior measure was the superior predictor.

**Table 2.**
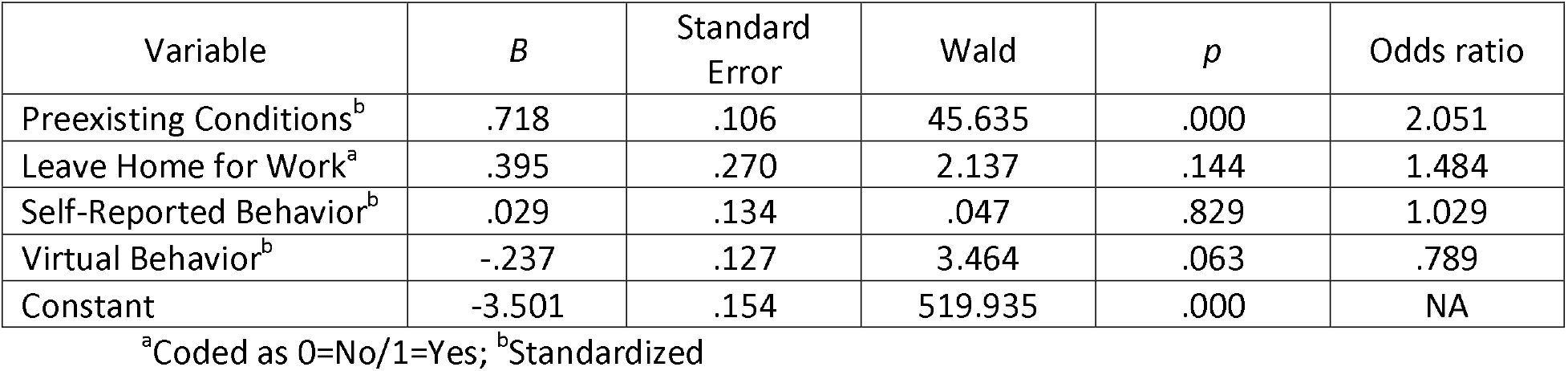
Binary Logistic Regression Predicting Having Tested Positively^a^.

## Discussion

The findings offer clear support regarding the efficacy of practicing social distancing. The more participants practiced social distancing, the less likely they were to have contracted COVID-19 over the next four months. In fact, an increase of one standard deviation on the virtual behavior measure of social distancing was associated with roughly a 20% reduction in the odds of contracting COVID-19. This was true both when considering only participants who reported having tested positively for the virus and when considering additional participants who, although untested, believed that they had contracted the virus.

Importantly, this prospective relation was observed at the level of the individual. The relation was apparent without aggregation across large numbers of people residing in a given locale – a characteristic of much of the research that has examined social distancing as a mitigating factor in the spread of the virus. In this respect, the current findings offer a unique form of additional evidence as to why individuals should indeed practice social distancing. What the individual does matters. It surely matters at the “collective”level, reducing transmission within the community – the very aim of officials’ social distancing directives and pleas. However, engaging in social distancing also carries with it clear and relatively immediate benefits for those who comply, leading to a reduced likelihood of contracting the disease for *that specific individual*.

Interestingly, however, our results also revealed a second critical contribution of the research. The virtual behavior measure of social distancing proved to be a better predictor of subsequent illness than did the self-report measure. The two measures were associated, but the correlation of .50 did not rise to a level that suggested these were essentially equivalent measures of the same construct, and differences in their predictive utility were evident. A significant relation was observed with the virtual measure both when predicting reports of believing one had contracted the virus and when restricting the sample to those who reported having tested positively. In contrast, the self-report measure revealed only a weak, marginally significant relation when predicting reports of illness and a null relation when predicting positive test outcomes. Moreover, when considered as simultaneous predictors, only the virtual distancing measure was associated with COVID-19 illness. These findings suggest a need for caution regarding the inferences that can be drawn from commonly-employed self-report measures of social distancing. If we had included only the self-report measure, the current study would have seriously underestimated the effectiveness of social distancing. Thus, the research findings also offer a methodological advance regarding the value of behavioral measures that simulate concrete, real-world decisions.

As noted earlier, self-report measures of behavior are open to a number of potential pitfalls. Respondents must reconstruct their past, a memory process that may itself be influenced by individuals’ beliefs about how they prefer to view themselves. Whatever is implied by this reconstructive memory process must then be aligned with the response options offered by the questionnaire item. For example, just what does following social distancing recommendations “very strictly” mean? To which subjective label or scale point does one’s assessment of the past correspond? Again, how individuals wish to view themselves or portray themselves to others is likely to affect how the scale points are disambiguated. These potential sources of bias are far less characteristic of the behavioral decisions required by the graphical scenarios that comprise our virtual behavior measure. Participants made concrete, “in-the-moment” decisions about alternative courses of action differing in the extent to which they allowed for social distancing. They interactively separated themselves visually from oncoming passersby; they selected a position on a crowded beach; they decided whether to walk along a route that required encountering other people. Participants made choices, much as they would in the real-world.

The data appear to suggest that a sizeable number of participants offered self-reports that were overestimates of their actual social distancing behavior. Whereas the virtual behavior data displayed a normal distribution, the distribution of scores on the self-report measure was skewed with a substantial majority responding at or near the positive endpoint of the scale (*M* = 5.98 on a 7-point scale, *SD* = 1.18). Such overestimation is to be expected to the extent that participants wished to believe themselves as having acted in manners that avoided placing their health, or that of others, at risk. Such self-beliefs have been shown to influence retrospective memory processes (e.g., Balcetis, 2008; Kouchaki & Gino, 2016; Ross, 1989).

Although the self-report measure was apparently “noisier” than the virtual behavior measure of social distancing, the current findings clearly illustrate the value of social distancing as a means of mitigating the likelihood of contracting COVID-19. The study offers *prospective* evidence to that effect. In contrast to past research, we show that social distancing matters not only at the aggregate level, but also critically at the level of the individual. Decisions that reflect the sound practice of social distancing reduce individuals’ risk of contracting COVID-19.

## Data Availability

The data are available on the Open Science Framework.

https://osf.io/x79ak/?view_only=b380d69aedc34bb98cf2670c04ecaa20

## Footnotes

1 See Fazio, Ruisch, Moore, Granados Samayoa, Boggs, & Ladanyi (2020) for a detailed report of Study 1.

2 Of these 1,686 cases, five involved missing values on one of the predictor variables. As a result, the analysis compared 199 cases of virus to 1,681 cases.

3 Once again, five of these latter cases involved missing values on one of the predictor variables. As a result, the analysis compared 85 positive test cases to 1,988 cases.

## References

Balcetis, E. (2008). Where the motivation resides and self-deception hides: How motivated cognition accomplishes self-deception. Social and Personality Psychology Compass, 2, 361–381.

Berinsky, A. J., Huber, G. A., & Lenz, G. S. (2012). Evaluating online labor markets for experimental research: Amazon.com’s Mechanical Turk. Political Analysis, 20, 351–368.

Buhrmester, M., Kwang, T., & Gosling, S. D. (2011). Amazon’s Mechanical Turk: A new source of inexpensive, yet high-quality, data? Perspectives on Psychological Science, 6, 3–5.

Chu, D. K., Akl, E.A., Duda, S., Yaacoub, S., Schünemann, H. J., et al. (2020). Physical distancing, face masks, and eye protection to prevent person-to-person transmission of SARS-CoV-2 and COVID-19: A systematic review and meta-analysis. The Lancet 2020 doi: 10.1016/S0140-6736(20)31142-9; 0810.1016/S0140-6736(20)31142-9.

Clifford, S., Jewell, R. M., & Waggoner, P. D. (2015). Are samples drawn from Mechanical Turk valid for research on political ideology? Research & Politics, 2. doi:10.1177/2053168015622072

Courtemanche, C., Garuccio, J., Le, A., Pinkston, J., & Yelowitz, A. (2020). Strong social distancing measures in the United States reduced the covid-19 growth rate. Health affairs (Project Hope), 39(7), 1237–1246. https://doi.org/10.1377/hlthaff.2020.00608

Fazio, R.H., Ruisch, B.C., Moore, C.A., Granados Samayoa, J.A., Boggs, S.T., & Ladanyi, J.T. (2020). Who is (not) complying with the social distancing directive and why? Testing a general framework of compliance with a behavioral measure of social distancing. medRxiv 2020.10.26.20219634; https://doi.org/10.1101/2020.10.26.20219634.

Ferguson, N. M., Laydon, D., Nedjati-Gilani, G., Imai, N., Ainslie, K., Baguelin, M., et al. (2020). Impact of non-pharmaceutical interventions (NPIs) to reduce COVID-19 mortality and healthcare demand. ImperialAcUk. 3–20. https://doi.org/10.25561/77482

Fisher, R. J. (1993). Social desirability bias and the validity of indirect questioning. Journal of Consumer Research, 20, 303–315.

Gollwitzer, A., Martel, C., Marshall, J., Höhs, J. M., & Bargh, J. A. (2020). Connecting self-reported social distancing to real-world behavior at the individual and U.S. state level. https://doi.org/10.31234/osf.io/kvnwp

Greenstone, M., & Nigam, V. (2020). Does social distancing matter? (March 30, 2020). University of Chicago, Becker Friedman Institute for Economics Working Paper No. 2020-26, Available at SSRN: http://dx.doi.org/10.2139/ssrn.3561244.

Gur, R. C., & Sackeim, H. A. (1979). Self-deception: A concept in search of a phenomenon. Journal of Personality and Social Psychology, 37, 147–169.

Hauser, D., Paolacci, G., & Chandler, J. (2019). Common concerns with MTurk as a participant pool Evidence and Solutions. In F. R. Kardes, P. M. Herr, & N. Schwarz (Eds.), Handbook of research methods in consumer psychology (pp. 319–336). New York, NY: Routledge.

Kissler, S.M., Tedijanto, C., Lipsitch, M., & Grad, Y. (2020). Social distancing strategies for curbing the COVID-19 epidemic. medRxiv. 2020.03.22.20041079.https://doi.org/10.1101/2020.03.22.20041079

Kouchaki, M., & Gino, F. (2016). Memories of unethical actions become obfuscated over time. Proceedings of the National Academy of Sciences, 113, 6166–6171.

Lee, E. C., Wada, N. I., Grabowski, M. K., Gurley, E. S., & Lessler, J. (2020). The engines of SARS-CoV-2 spread. Science, 370 (6515), 406–407.

Leary, M. R., & Kowalski, R. M. (1990). Impression management: A literature review and two component model. Psychological Bulletin, 107, 34–47.

McGrail, D. J., Dai, J., McAndrews, K. M., & Kalluri, R. (2020). Enacting national social distancing policies corresponds with dramatic reduction in COVID19 infection rates. PLoS ONE 15(7): e0236619. https://doi.org/10.1371/journal.pone.0236619

Paolacci, G., & Chandler, J. (2014). Inside the Turk: Understanding mechanical turk as a participant pool. Current Directions in Psychological Science, 23, 184–188.

Ross, M. (1989). Relation of implicit theories to the construction of personal histories. Psychological Review, 96, 341–357.

VoPham, T., Weaver, M. D., Hart, J. E., Ton, M., White, E., & Newcomb, P. A. (2020). Effect of social distancing on COVID-19 incidence and mortality in the US. medRxiv 2020.06.10.20127589; https://doi.org/10.1101/2020.06.10.20127589

